# Temporal Feature Engineering and Ensemble Learning for Predicting 28-Day Mortality in ICU Patients with Alcoholic Cirrhosis

**DOI:** 10.64898/2026.06.30.26356958

**Authors:** Janet Sanjaya, Mohammadsaeed Haghi, Nausin Kudrot, Sakshie Pathak, Shreyas V. Chandramouli, Kamiar Alaei, Maryam Pishgar

## Abstract

**Background:** Predicting 28-day mortality in ICU patients with alcoholic cirrhosis is challenging because clinical deterioration is dynamic and heterogeneous.

**Methods:** Using MIMIC-IV (v3.1), this study included 1,907 patients (training ***n* = 1**,**334**; validation ***n* = 573**), engineering 208 temporal and static predictors from 64 base variables and reducing them to 40 through multi-stage selection. Seven classifiers and a weighted gradient-boosting ensemble (XGBoost, CatBoost, LightGBM) were compared with Optuna tuning.

**Results:** The ensemble achieved the highest internal validation AUC (0.9276; 95% CI: 0.9011–0.9507) and lowest Brier score (0.0870), with strong discrimination on eICU-CRD (AUC 0.9347) and related MIMIC-III (AUC 0.9071). Ablation indicated that temporal features, especially deltas, were major contributors (**Δ**AUC **≈ 0.17** when removed). SHAP highlighted APS III score, anion gap, oxygen saturation (delta), lactate, and INR as leading predictors.

**Conclusions:** The framework supports interpretable, trajectory-informed risk stratification in critically ill cirrhotic patients; prospective validation is needed before clinical use.

## 1 Background

Accurate prediction of patient outcomes remains a central challenge in intensive care, where heterogeneous conditions, nonlinear physiological interactions, and evolving clinical states make mortality difficult to forecast. Among critically ill patients, those with alcoholic cirrhosis are particularly vulnerable, combining progressive liver dysfunction with systemic complications and high short-term mortality. Alcohol-related liver disease is a major contributor to cirrhosis mortality in the United States [1–3], and reported ICU mortality for these patients ranges from 36% to 65%, underscoring the need for reliable risk stratification.

Traditional severity scores such as the Model for End-Stage Liver Disease (MELD) are widely used to estimate mortality risk in cirrhosis, but they rely on a small set of static laboratory variables and were designed for organ allocation rather than ICU prognosis. Their discrimination is correspondingly moderate, with reported area under the curve (AUC) values around 0.77 [4, 5], and they do not capture temporal patterns or nonlinear interactions that characterize critically ill patients.

Machine learning has been increasingly applied to electronic health record data for ICU mortality prediction, and the availability of large databases such as MIMIC-IV [6, 7] has accelerated this work. In particular, tree-based and boosting methods have outperformed conventional scores [8–16]. A prior MIMIC-IV study [13] predicted 28-day mortality in alcoholic cirrhosis using logistic regression, decision trees, and gradient boosting, but relied primarily on static features. Related work has addressed short-term mortality in critically ill cirrhotic patients [12], cirrhosis with acute kidney injury [14], and dynamic or explainable ICU models more broadly [9, 11, 15, 16].

Despite this progress, important gaps remain. Most models in this setting still underuse temporal information, such as trends and changes in vitals and laboratory values, that may better reflect ICU deterioration than static admission measurements. In addition, studies often lack systematic comparison across model families, calibration assessment, interpretability analysis, and validation on independent cohorts.

To address these gaps, this study developed a machine learning framework for 28-day all-cause mortality in ICU patients with alcoholic cirrhosis using MIMIC-IV (v3.1). Trajectory-based predictors were engineered from longitudinal vitals and laboratories, applied multi-stage feature selection, compared seven learners and a weighted gradient-boosting ensemble with Optuna tuning, and evaluated discrimination, calibration, SHAP interpretability, and ablation analyses, followed by frozen-model evaluation on eICU-CRD and a related MIMIC-III cohort. The main contributions are: (1) temporal feature engineering from 64 base variables into means, deltas, slopes, and missingness indicators; (2) a reproducible selection pipeline (filter, embedded ranking, RFECV, and post-tuning leave-one-out ablation) yielding 40 predictors; (3) systematic benchmarking of seven algorithms and a weighted ensemble; and (4) external and related-cohort evaluation to assess transportability beyond the development sample. Reporting followed the Transparent Reporting of a multivariable prediction model for Individual Prognosis Or Diagnosis (TRIPOD) guidelines [17, 18].

## 2 Methods

### 2.1 Data source and ethics

This study used the Medical Information Mart for Intensive Care IV (MIMIC-IV, version 3.1), a publicly available electronic health record database hosted on PhysioNet [6, 7, 19]. MIMIC-IV contains de-identified health-related data for patients admitted to the intensive care units of Beth Israel Deaconess Medical Center (Boston, Massachusetts) between 2008 and 2019. The database integrates hospital, ICU, and derived tables, including demographics, admissions, diagnoses, procedures, laboratory results, vital signs, and summary clinical indicators (Figure 1).

**Fig. 1.**
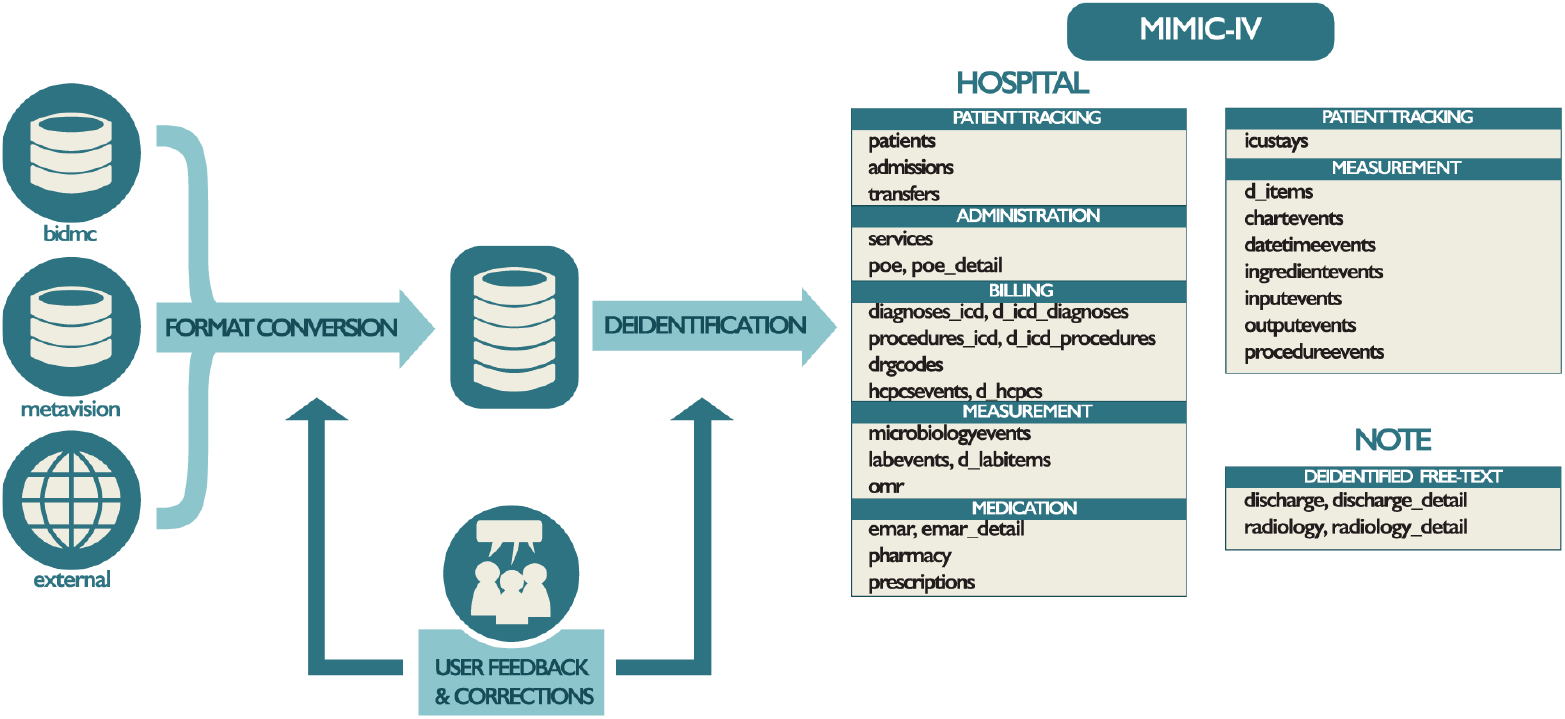
Overview of the MIMIC-IV data acquisition and database structure. Adapted from Johnson et al. [6].

As shown in Figure 1, MIMIC-IV links data from multiple hospital information systems into a structured relational format. Both static and longitudinal measurements are available, supporting extraction of time-dependent predictors for temporal feature engineering (Section 2.4).

Data were extracted with structured query language (SQL) in Google BigQuery. Records were retrieved from hospital-level, ICU-level, and derived tables and merged at the patient–ICU-stay level for cohort construction and feature engineering.

Access to MIMIC-IV requires credentialed registration and completion of required training on PhysioNet. Because the database is de-identified, institutional review board approval was not required for this secondary analysis of existing data.

### 2.2 Study population and cohort selection

Adult ICU patients with alcoholic cirrhosis were identified from discharge diagnoses using International Classification of Diseases (ICD) codes ICD-9 571.2 and ICD-10 K70.30 and K70.31. Exclusion criteria were age < 18 years, ICU length of stay < 24 hours, and duplicate admissions (only the first eligible ICU stay per patient was retained).

Of 3,531 initially identified records, 1,624 were excluded, yielding 1,907 patients. Within 28 days of ICU admission, 473 patients (24.8%) died and 1,434 (75.2%) survived, indicating a moderately imbalanced outcome distribution. The cohort was divided into training (*n* = 1,334, 70%) and internal validation (*n* = 573, 30%) sets using stratified random sampling on 28-day mortality, so the outcome prevalence was preserved in both subsets (Figure 2).

**Fig. 2.**
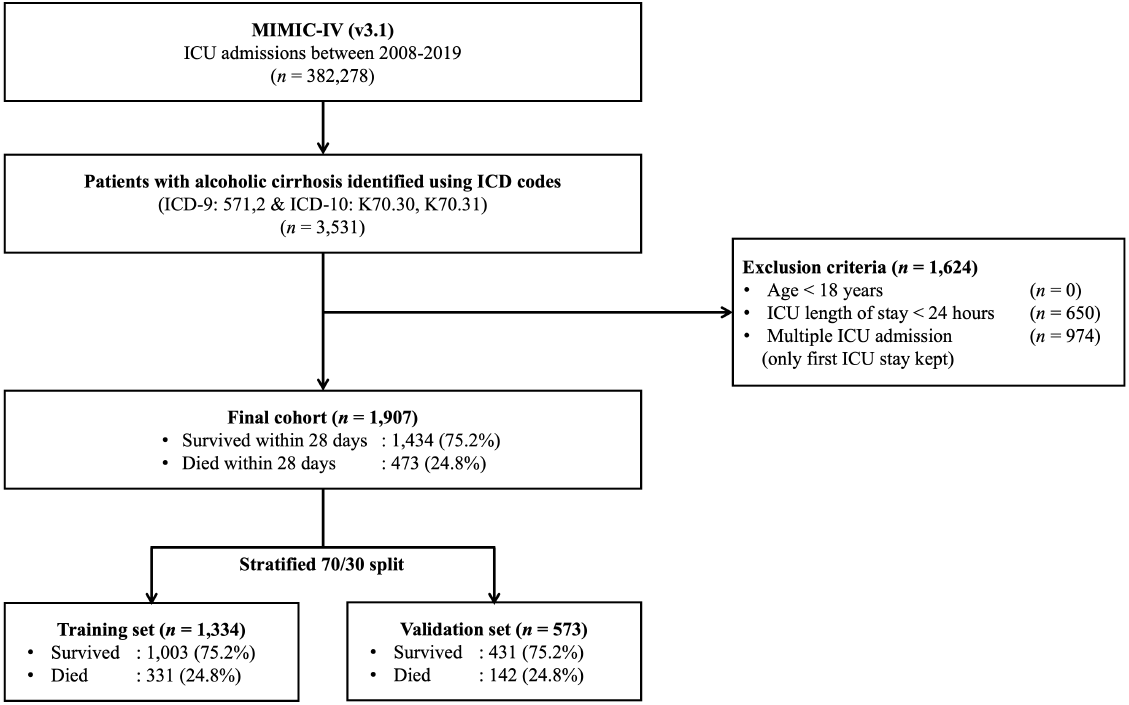
Flowchart of cohort selection and dataset construction for ICU patients with alcoholic cirrhosis.

Baseline (non-temporal) characteristics of the training and validation groups were compared descriptively (Table 4). Continuous variables are reported as mean ± standard deviation and categorical variables as count (%). Group comparisons used the Mann–Whitney *U* test for continuous variables and the chi-square or Fisher’s exact test for categorical variables, as appropriate.

**Table 1.** List of candidate features used in the study.

| Category | Variables |
| --- | --- |
| **Demographics** | Age, BMI, Gender, Insurance, Race |
| **Disease Severity Scores** | APS III score, LODS score, MELD-NA score, OASIS score, SAPS II score, SOFA score |
| **Comorbid Conditions** | Atrial fibrillation, Chronic kidney disease, Chronic obstructive pulmonary disease, Congenital coagulation defects, Coronary artery disease, Heart failure, Malignant tumor, Sepsis, Stroke |
| **Liver-related Conditions** | Ascites, Hepatic encephalopathy, Hepatorenal syndrome, Portal hypertension, Spontaneous bacterial peritonitis, Variceal bleeding |
| **Vital Signs** | Diastolic blood pressure, Heart rate, Mean arterial pressure, Oxygen saturation, Respiratory rate, Systolic blood pressure, Temperature |
| **Laboratory Measurements** | Activated partial thromboplastin time (aPTT), Albumin, Alkaline phosphatase, Alanine aminotransferase (ALT), Anion gap, Aspartate aminotransferase (AST), Bilirubin, Calcium, Chloride, Creatinine, C-reactive protein (CRP), Estimated glomerular filtration rate (eGFR), Fibrinogen, Fraction of inspired oxygen (FiO<sub>2</sub>), Glucose, Hematocrit, Hemoglobin, International normalized ratio (INR), Lactate, Magnesium, Neutrophil count, Partial pressure of oxygen (PaO<sub>2</sub>), PF (PaO<sub>2</sub>/FiO<sub>2</sub>) ratio, Phosphate, Platelet count, Potassium, Prothrombin time, Total protein, White blood cell count |
| **Interventions** | Mechanical ventilation, Vasopressor |

**Table 2.**
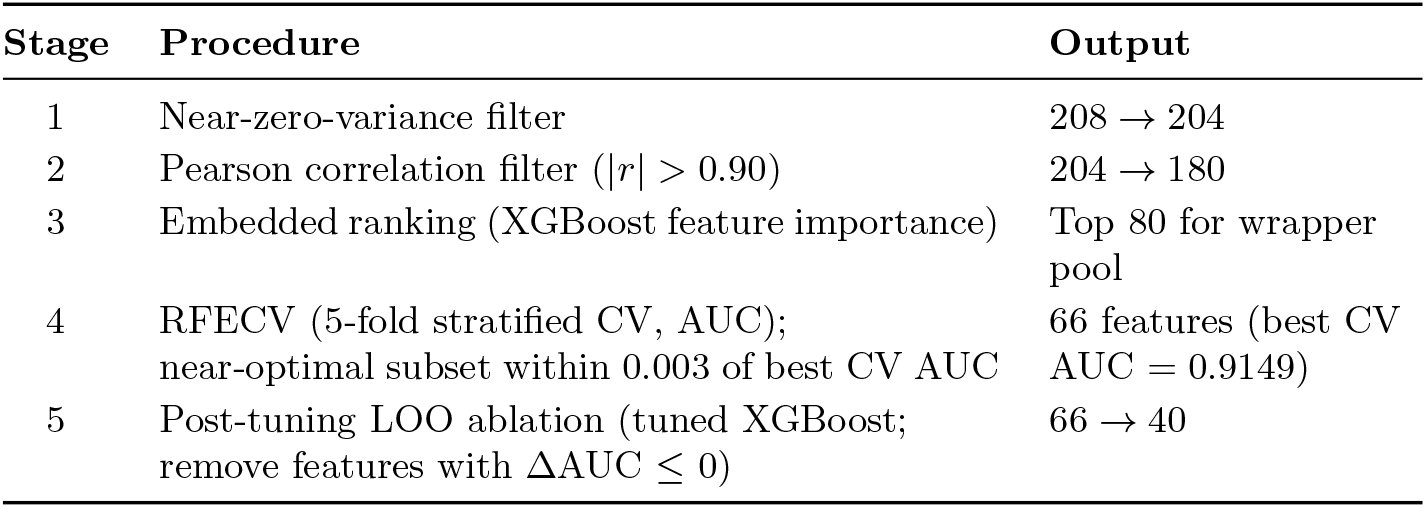
Multi-stage feature selection pipeline applied to the training split.

**Table 3.** Variables removed by post-tuning leave-one-out (LOO) ablation on the tuned XGBoost model.

| Removed variable | LOO validation impact | Rationale |
| --- | --- | --- |
| Activated partial thromboplastin time (baseline) | Validation AUC higher when removed ($\Delta\text{AUC} = -0.0032$) | Adverse/redundant contribution |
| Lactate (missing indicator) | Validation AUC higher when removed ($\Delta\text{AUC} = -0.0031$) | Adverse/redundant contribution |
| Oxygen saturation (baseline) | Validation AUC higher when removed ($\Delta\text{AUC} = -0.0030$) | Adverse/redundant contribution |
| Activated partial thromboplastin time (delta) | Validation AUC higher when removed ($\Delta\text{AUC} = -0.0026$) | Adverse/redundant contribution |
| Anion gap (slope) | Validation AUC higher when removed ($\Delta\text{AUC} = -0.0024$) | Adverse/redundant contribution |
| Creatinine (delta) | Validation AUC higher when removed ($\Delta\text{AUC} = -0.0023$) | Adverse/redundant contribution |
| Alanine aminotransferase (mean) | Validation AUC higher when removed ($\Delta\text{AUC} = -0.0022$) | Adverse/redundant contribution |
| Glucose (slope) | Validation AUC higher when removed ($\Delta\text{AUC} = -0.0022$) | Adverse/redundant contribution |
| Chloride (mean) | Validation AUC higher when removed ($\Delta\text{AUC} = -0.0021$) | Adverse/redundant contribution |
| White blood cell count (slope) | Validation AUC higher when removed ($\Delta\text{AUC} = -0.0021$) | Adverse/redundant contribution |
| Partial pressure of oxygen (slope) | Validation AUC higher when removed ($\Delta\text{AUC} = -0.0015$) | Adverse/redundant contribution |
| Temperature (baseline) | Validation AUC higher when removed ($\Delta\text{AUC} = -0.0015$) | Adverse/redundant contribution |
| Aspartate aminotransferase (delta) | Validation AUC higher when removed ($\Delta\text{AUC} = -0.0014$) | Adverse/redundant contribution |
| Creatinine (slope) | Validation AUC higher when removed ($\Delta\text{AUC} = -0.0014$) | Adverse/redundant contribution |
| Respiratory rate (slope) | Validation AUC higher when removed ($\Delta\text{AUC} = -0.0014$) | Adverse/redundant contribution |

Table 3 – continued
| Removed variable | LOO validation impact | Rationale |
| --- | --- | --- |
| Magnesium (delta) | Validation AUC higher when removed ($\Delta\text{AUC} = -0.0011$) | Adverse/redundant contribution |
| Mean arterial pressure (mean) | Neutral or slight improvement ($\Delta\text{AUC} = -0.0010$) | Removed for parsimony |
| International normalized ratio (slope) | Neutral or slight improvement ($\Delta\text{AUC} = -0.0009$) | Removed for parsimony |
| Lactate (slope) | Neutral or slight improvement ($\Delta\text{AUC} = -0.0008$) | Removed for parsimony |
| Oxygen saturation (mean) | Neutral or slight improvement ($\Delta\text{AUC} = -0.0008$) | Removed for parsimony |
| Prothrombin time (delta) | Neutral or slight improvement ($\Delta\text{AUC} = -0.0005$) | Removed for parsimony |
| Diastolic blood pressure (mean) | Neutral or slight improvement ($\Delta\text{AUC} = -0.0003$) | Removed for parsimony |
| Mean arterial pressure (delta) | Neutral or slight improvement ($\Delta\text{AUC} = -0.0003$) | Removed for parsimony |
| Fibrinogen (slope) | Neutral or slight improvement ($\Delta\text{AUC} = -0.0002$) | Removed for parsimony |
| Phosphate (delta) | Neutral or slight improvement ($\Delta\text{AUC} = -0.0002$) | Removed for parsimony |
| Glucose (baseline) | Negligible impact ($\Delta\text{AUC} \approx 0$) | Removed for parsimony |

**Table 4.**
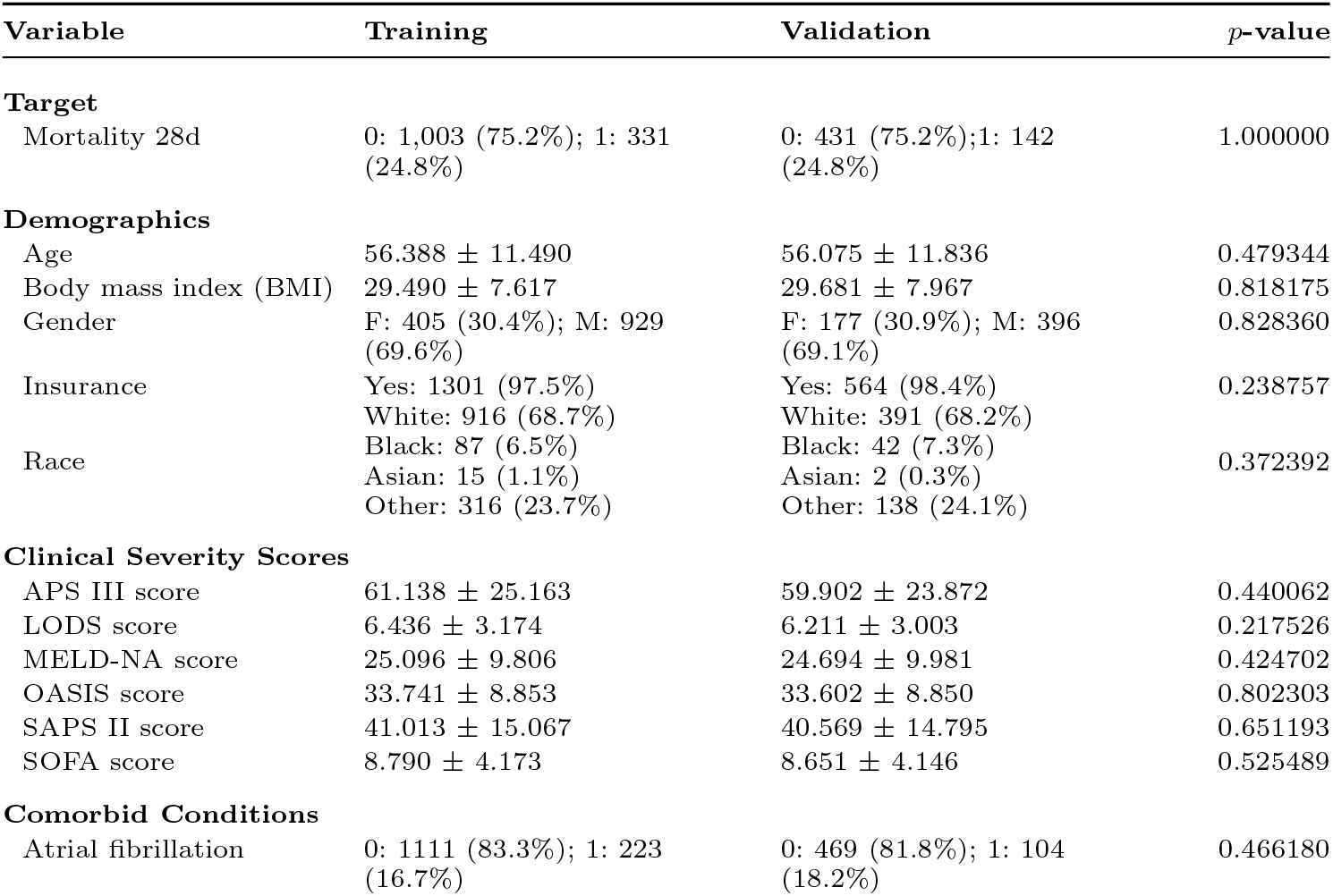
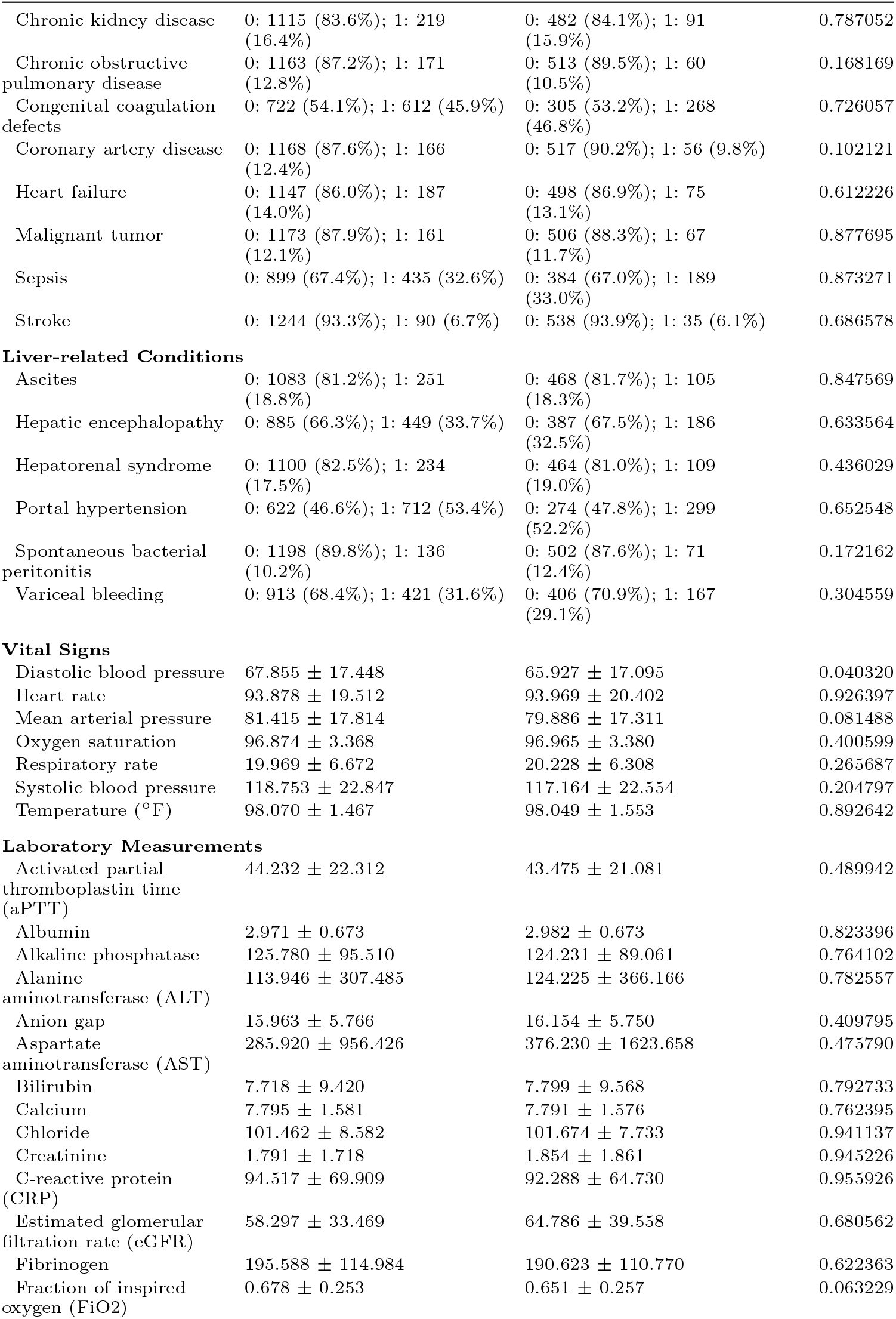
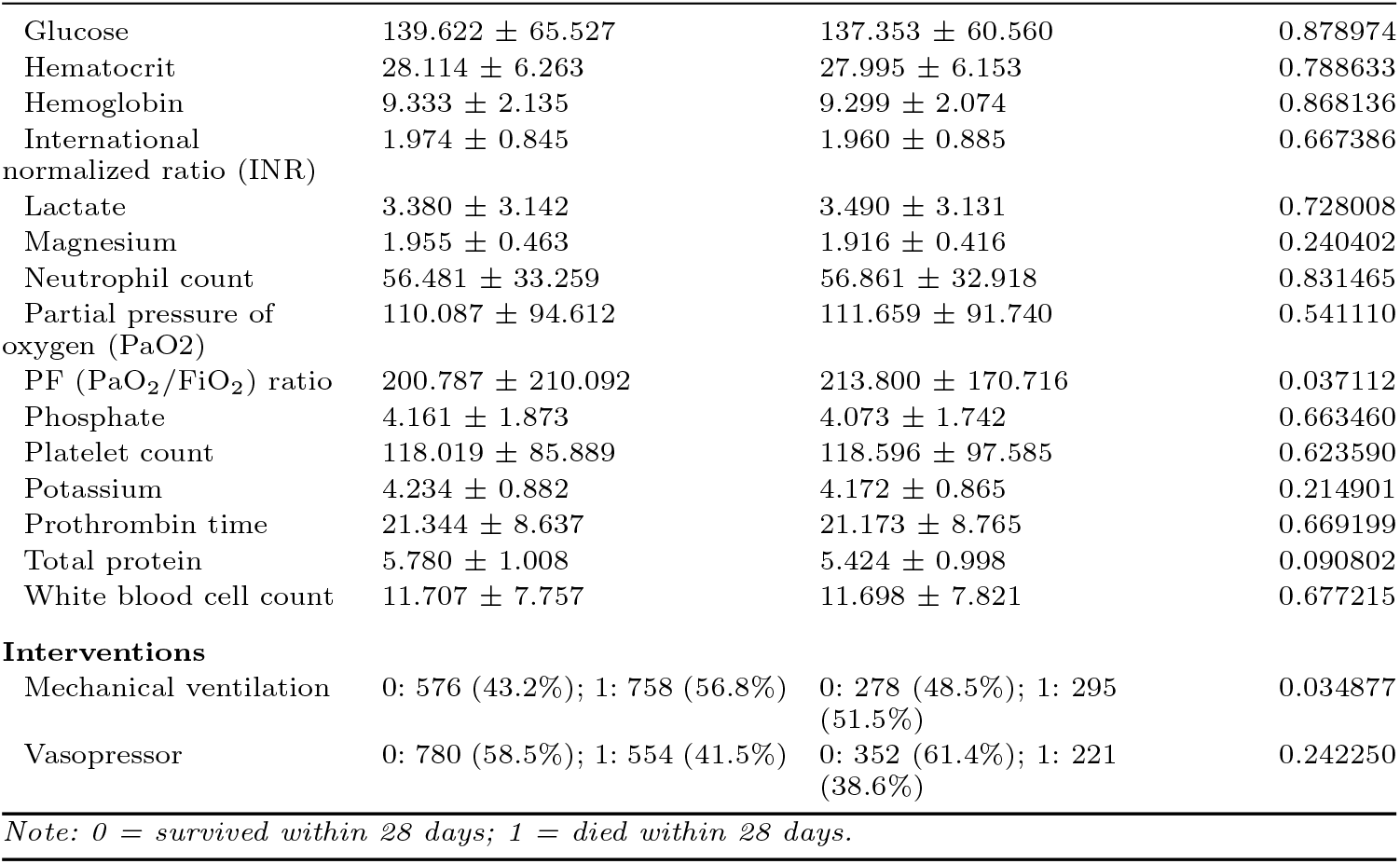
Baseline characteristics of the training and validation cohorts.

### 2.3 Outcome definition

The primary outcome was 28-day all-cause mortality, defined as death from any cause within 28 days of ICU admission and coded as a binary variable (1 = death, 0 = survival). Outcome status was determined by comparing recorded death dates with ICU admission time in MIMIC-IV.

### 2.4 Predictor variables and temporal feature construction

Predictors were extracted from MIMIC-IV and grouped into demographics, comorbidities, liver-related complications, severity scores, vital signs, laboratory measurements, and interventions (Table 1). Time-dependent laboratory and vital-sign variables were expanded into trajectory summaries; static variables (demographics, severity scores, comorbidities, liver-related conditions, and interventions) were retained without transformation.

For a longitudinal variable with values *x*_1_, …, *x*_*n*_ recorded at times *t*_1_, …, *t*_*n*_ during the ICU stay (*t* measured from ICU admission), the engineered representations were

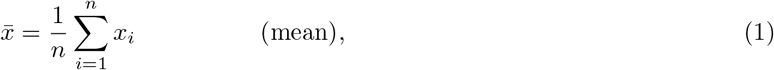

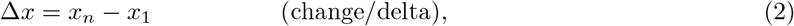

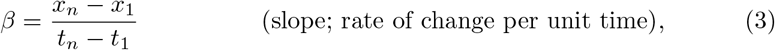

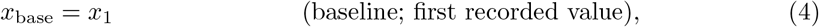

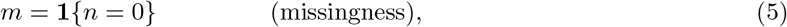

where *x*_1_ and *x*_*n*_ are the first and last observed values, *t*_1_ and *t*_*n*_ the corresponding timestamps, and **1**{·} the indicator function. Timestamps were converted to hours, so *β* is the average hourly rate of change between the first and last measurements. For each time-dependent variable, the baseline value was stored without a suffix (e.g., `lactate`), and derived columns were appended as `_mean, _delta, _slope`, and `_missing`. This expanded 64 base clinical variables into 208 candidate predictors (Algorithm 1).

#### Algorithm 1 Temporal feature construction for time-dependent predictors

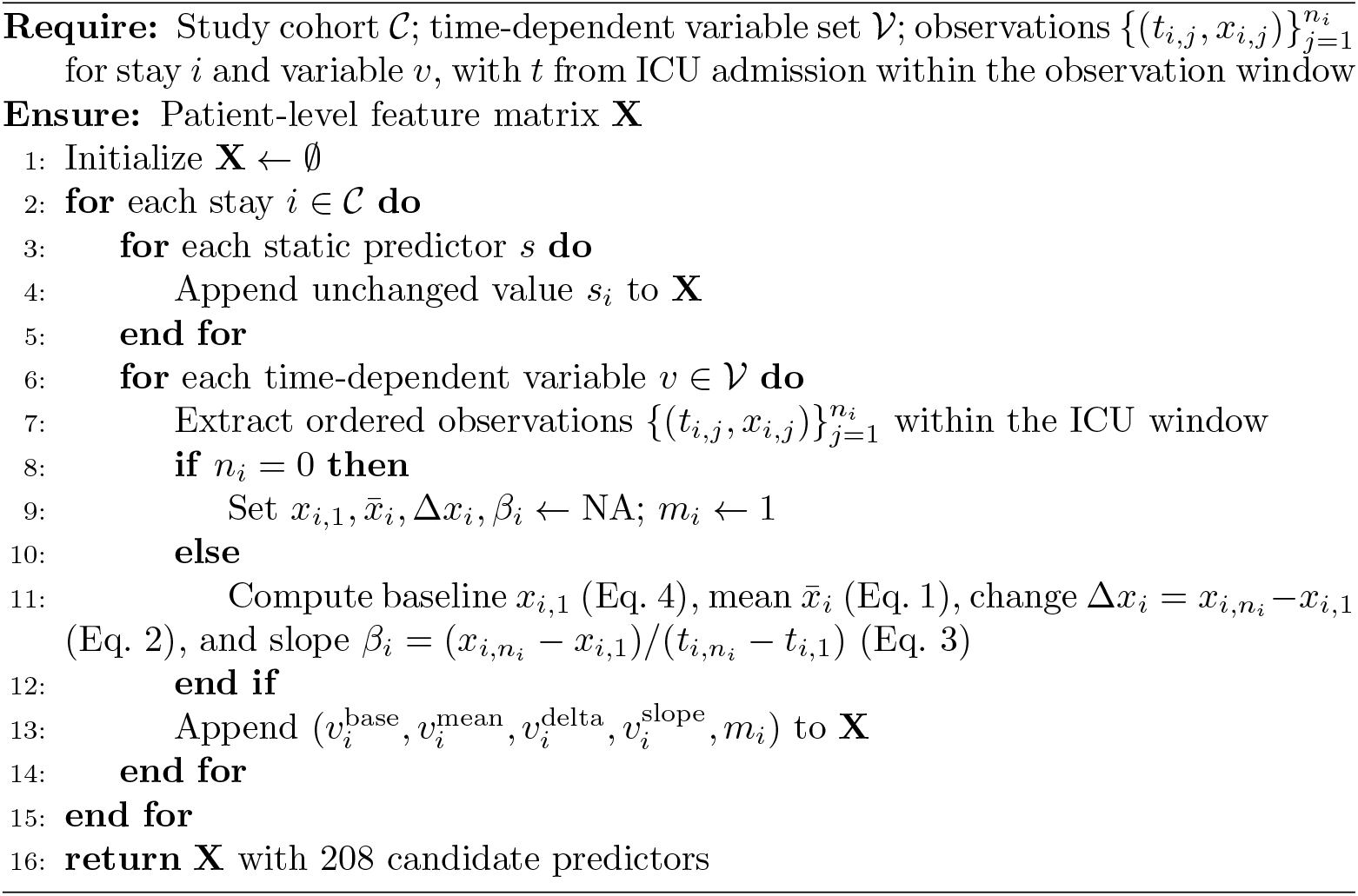

### 2.5 Data preprocessing

All preprocessing steps were fit on the training split only and applied unchanged to the internal validation and external cohorts to prevent data leakage. Implausible baseline and mean values were set to missing using predefined clinical rules (e.g., out-of-range mean arterial pressure, SpO_2_, heart rate, respiratory rate, FiO_2_, temperature, and prothrombin time; temperature in degrees Celsius was converted to Fahrenheit before range checks). Calcium values below 4 mmol/L were multiplied by 4.0 to harmonize mixed units; glucose values exceeding 1000 mg/dL were set to missing. Delta and slope features were not re-clipped.

Categorical variables (gender, insurance, race) were encoded numerically: gender and insurance as binary variables; race standardized to White, Black, Asian, and Other with one-hot encoding and one reference category omitted. Non-numeric values were coerced to missing; features entirely missing in the training set were dropped. Remaining missing numeric values were imputed with training-set medians, which are robust to outliers and appropriate for skewed clinical distributions. Standardization (training mean and standard deviation) was applied only for logistic regression and SVM during internal development; tree-based models, including the final ensemble members, were trained on imputed, unscaled features.

Class imbalance was addressed during training with XGBoost `scale_pos_weight` set to the training-set negative-to-positive ratio; logistic regression, SVM, decision tree, random forest, and LightGBM used `class_weight = “balanced”`; CatBoost used no additional class-weight specification. All imbalance adjustments were computed from training-set class counts only.

### 2.6 Feature selection

Following temporal feature construction, the modeling dataset contained 208 candidate features derived from 64 base clinical variables. To reduce dimensionality and improve model efficiency while preserving predictive performance, a multi-stage feature selection strategy was applied on the training split only, consisting of filter, embedded, and wrapper methods (Table 2).

Filter methods were applied first to remove low-information and redundant predictors. Near-zero-variance features were eliminated (208 → 204). Highly correlated variables were then removed using a Pearson correlation threshold of |*r*| > 0.90, with correlations estimated on the training set only. This step reduced redundancy and multicollinearity and yielded 180 features.

Next, an embedded ranking was performed with an Extreme Gradient Boosting (XGBoost) model trained on the filtered set with class-imbalance weighting. Feature importance scores were used to rank predictors, leveraging the ability of tree-based models to capture nonlinear relationships and interactions in clinical data. The top 80 ranked features formed the wrapper pool, balancing computational feasibility with feature diversity.

Wrapper selection used recursive feature elimination with cross-validation (RFECV) on this pool. RFECV iteratively removed features and evaluated performance using 5-fold stratified cross-validated AUC. Let *S*_feas_ denote the feature subsets evaluated during RFECV and AUC_cv_(*S*) the corresponding cross-validated AUC. Rather than selecting the subset with the absolute highest AUC, the smallest near-optimal subset was retained:

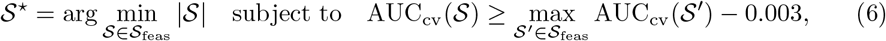

yielding |*S*^⋆^| = 66 features (best RFECV cross-validated AUC 0.9149).

Hyperparameter tuning (Section 2.8) was performed on the 66-feature set. Post-tuning leave-one-out (LOO) ablation was then applied to the tuned XGBoost model. For each feature *j*, the change in internal validation AUC after removing *j* alone was defined as

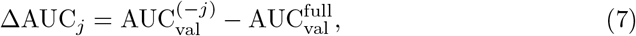

where 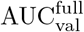 is validation AUC with all 66 predictors and 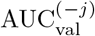 is validation AUC after removing feature *j*. Features with **Δ**AUC_*j*_ ≤ 0 were excluded, removing 26 variables and retaining 40 predictors (Table 3; final list in Table 5 in Results). Unlike hyperparameter and ensemble-weight optimization, which used training-set cross-validation only, LOO exclusion used internal validation AUC and therefore constitutes hold-out information use during predictor reduction.

**Table 5.**
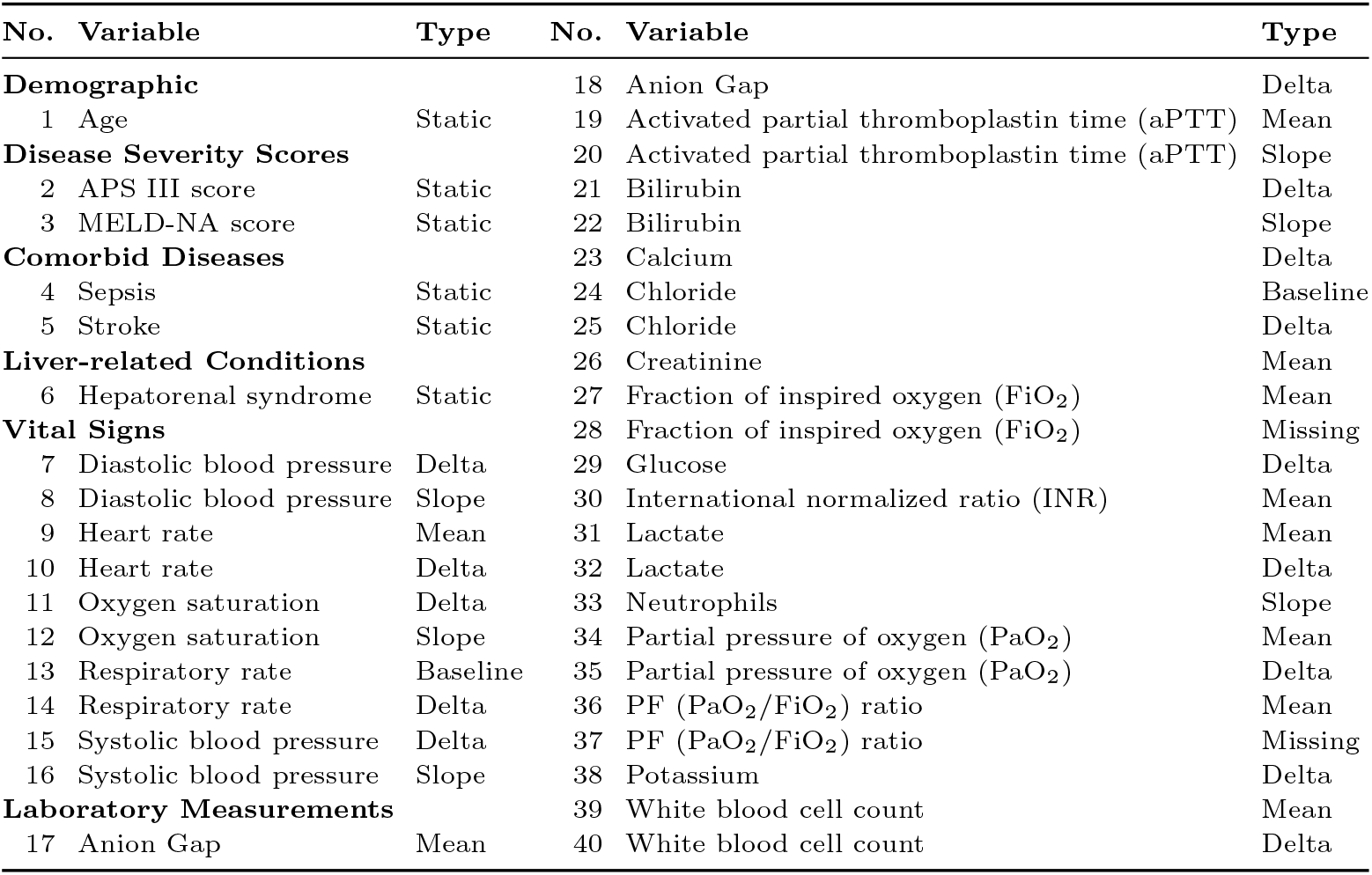
Final 40 features used for model development, internal validation, and external validation.

### 2.7 Machine learning models and ensemble

Seven classifiers were compared: logistic regression, support vector machine (SVM) with radial basis function (RBF) kernel, decision tree, random forest [20], XGBoost [21], LightGBM [22], and CatBoost [23]. Logistic regression served as an interpretable linear baseline; SVM with an RBF kernel captured nonlinear boundaries; decision tree and random forest represented single-tree and bagged ensemble approaches; and the three gradient-boosting models were included because they commonly perform strongly on structured clinical tabular data and can model nonlinear effects and feature interactions. All models output predicted 28-day mortality probabilities, enabling systematic comparison of linear, kernel-based, and tree-based learners on the same predictor set and tuning protocol.

Hyperparameters for each learner were optimized with Optuna on the training split (Section 2.8); fixed parameter values are not repeated here. The primary reported model was a weighted ensemble of XGBoost, CatBoost, and LightGBM. For ensemble member *m* ∈ {XGB, CAT, LGBM}, let 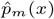 denote the predicted 28-day mortality probability for feature vector *x*. The ensemble probability was computed as

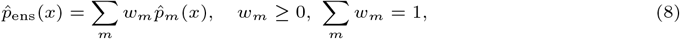

where nonnegative weights *w*_*m*_ were optimized on training-set out-of-fold probabilities (Section 2.8). This weighted average leverages complementary boosting approaches and improves stability relative to any single model.

Because models output predicted mortality probabilities, binary high-risk classification requires an operating threshold. A default cutoff of 0.5 is often inappropriate when mortality is moderately uncommon and missed deaths carry high clinical cost. High-risk classification was defined as

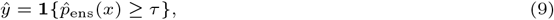

where *τ* was selected on the internal validation set for the final 40-feature weighted ensemble from a grid of 150 equally spaced values on [0.05, 0.90] by maximizing

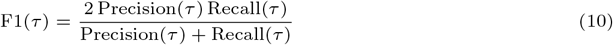

subject to Recall(*τ* ) ≥ 0.80, yielding *τ* = 0.1013. Thus, patients with estimated 28-day mortality probability of at least 10.1% were classified as high risk. This rule prioritized sensitivity while limiting false alarms among thresholds meeting the minimum recall requirement. The selected threshold was frozen and applied unchanged in external evaluation.

### 2.8 Hyperparameter tuning

Hyperparameters were optimized with Optuna [24] using the Tree-structured Parzen Estimator (TPE) sampler (40 trials per study). This Bayesian optimization approach enables efficient exploration of the hyperparameter space compared with traditional grid search or random search, while maintaining strong performance in high-dimensional search problems with limited trial budgets.

Each base model (logistic regression, SVM, decision tree, random forest, XGBoost, LightGBM, and CatBoost) was tuned on the 66-feature RFECV predictor set derived from the training split. Candidate configurations were evaluated with repeated stratified 5-fold cross-validation (3 repeats) on the training data. Stratification preserved the 28-day mortality prevalence in every fold, which is important given the moderately imbalanced outcome and helps prevent folds with unusually few deaths that would make cross-validated AUC unstable. Repeating the 5-fold procedure reduced sensitivity to a single random partition and provided a more stable estimate of out-of-sample discrimination. The hyperparameter configuration with the highest mean cross-validated AUC was selected for each model. Random seed 42 was fixed for data splitting, cross-validation, and stochastic algorithms to support reproducibility.

Ensemble weights were optimized in a separate Optuna study (40 trials) on the same 66-feature set. For each gradient-boosting member (XGBoost, CatBoost, and LightGBM), out-of-fold (OOF) predicted probabilities were generated on the training set using a single stratified 5-fold pass (without the 3-repeat scheme used for base-model tuning). Within each trial, nonnegative member weights were assigned, normalized to sum to 1, and combined into a weighted ensemble probability; the objective was OOF ROC-AUC on training data. The weight configuration with the highest OOF AUC was selected.

After tuning, LOO ablation reduced the predictor set from 66 to 40 features; tuned hyperparameters and ensemble weights were retained without re-optimization on the reduced set. Models were then refit on the full training split using the 40-feature matrix. Hyperparameter and weight selection used training data only; the internal validation split was reserved for LOO exclusion, threshold selection, and final internal performance reporting. The frozen export bundle comprised the 40-feature list, training-fit median imputer, refitted tree-based estimators, ensemble weights, and classification threshold.

Algorithm 2 summarizes the end-to-end development workflow and information-use boundaries across training, validation, and external cohorts.

#### Algorithm 2 Model development, selection, and frozen external validation workflow

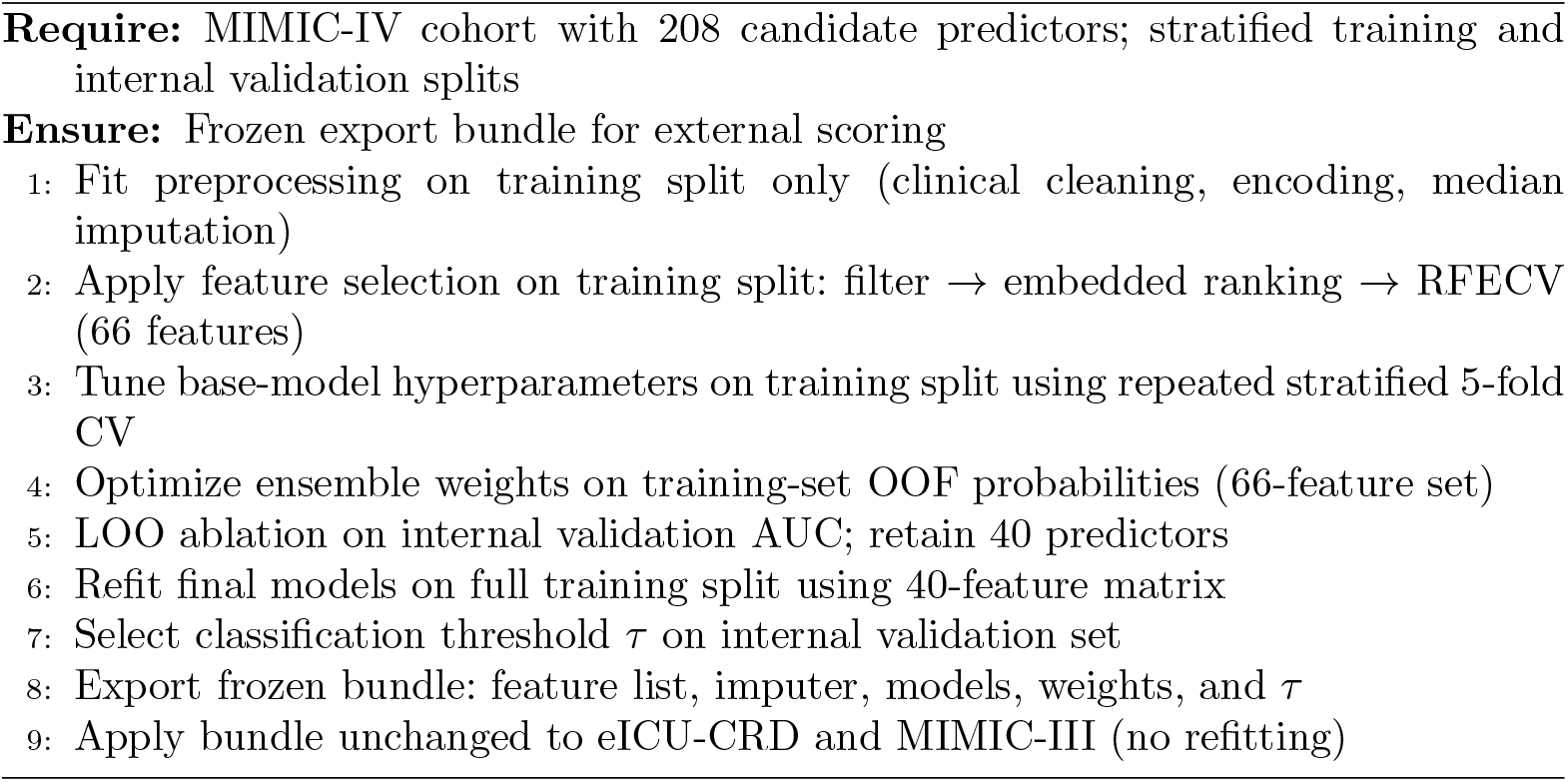

### 2.9 Model evaluation and statistical analysis

Discrimination was assessed primarily with the area under the receiver operating characteristic curve (AUC). At the validation-selected threshold, precision, recall (sensitivity), F1-score, and confusion matrices were reported. AUC uncertainty was estimated with 1,000 bootstrap replicates on each evaluation cohort.

Calibration was assessed with calibration curves and the Brier score:

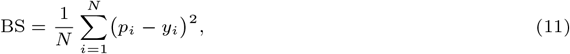

where *p*_*i*_ is the predicted probability and *y*_*i*_ ∈ {0, 1} the observed outcome for patient *i*.

Interpretability was examined with SHAP (Shapley Additive Explanations) on tree-based ensemble members. For prediction *f* (*x*) with feature vector *x* = (*x*_1_, …, *x*_*d*_), SHAP decomposes 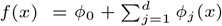, where *ϕ*_0_ is the expected output over the training distribution and *ϕ*_*j*_ (*x*) quantifies the contribution of feature *x*_*j*_ .

Robustness was examined with post-hoc ablation on the tuned XGBoost model using the final 40-feature set: feature-group ablation (laboratory, vital signs, comorbidities, liver-related conditions, and severity scores) and temporal ablation (temporal vs. non-temporal predictors and representation types). Group-level and temporal ablation results are reported in Results (Figure 7); these analyses quantified contribution to discrimination and did not further reduce the final predictor set.

**Fig. 3.**
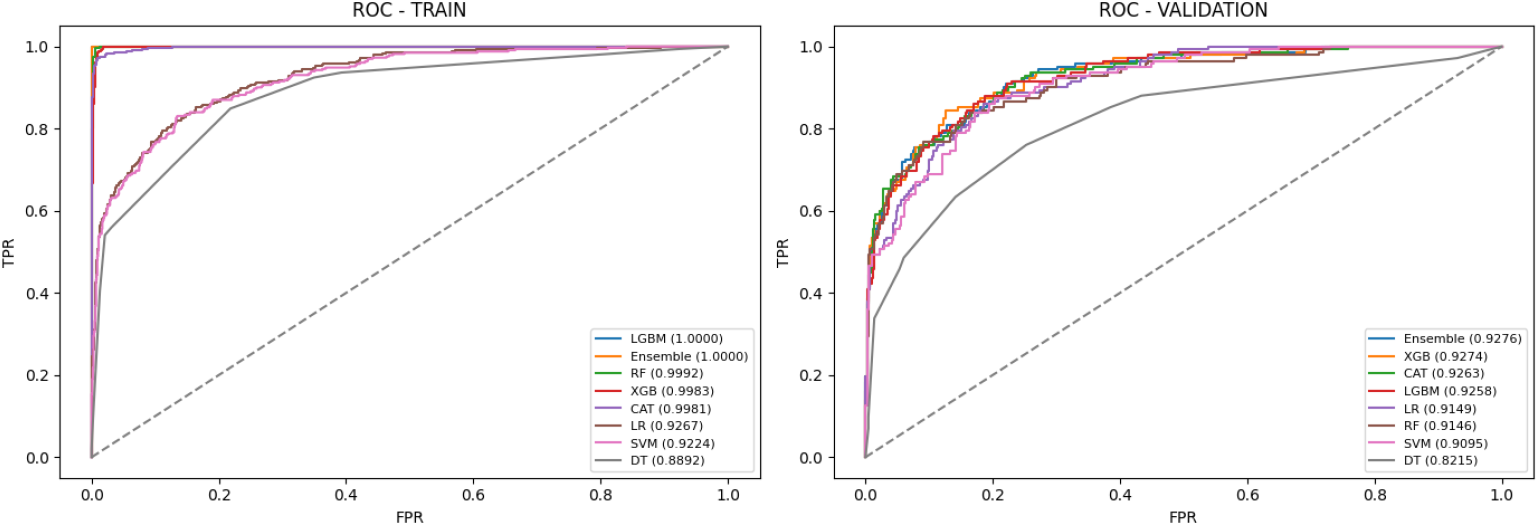
ROC curves of the evaluated models on the training and validation sets.

**Fig. 4.**
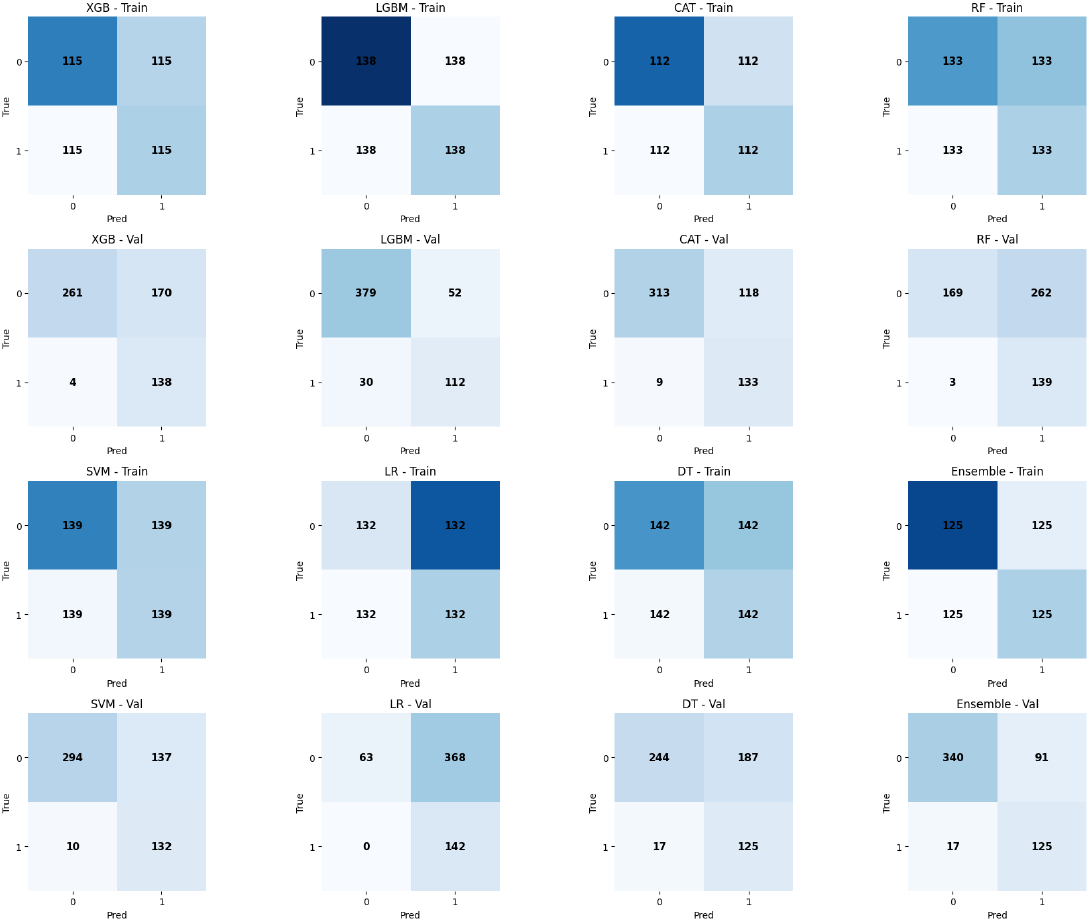
Confusion matrices on the training and validation sets at threshold *τ* = 0.1013.

**Fig. 5.**
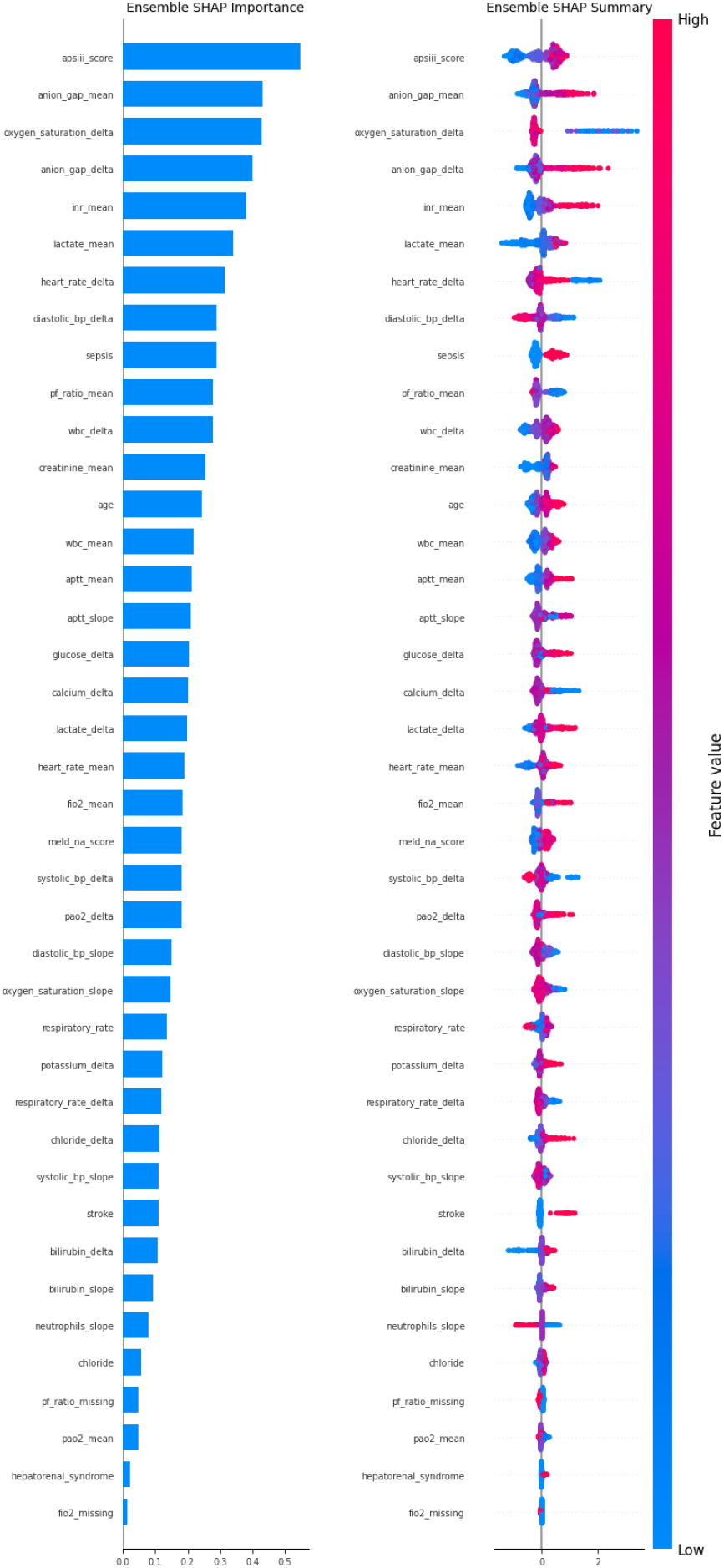
SHAP feature importance (bar and beeswarm plot) for the ensemble model on the validation set.

**Fig. 6.**
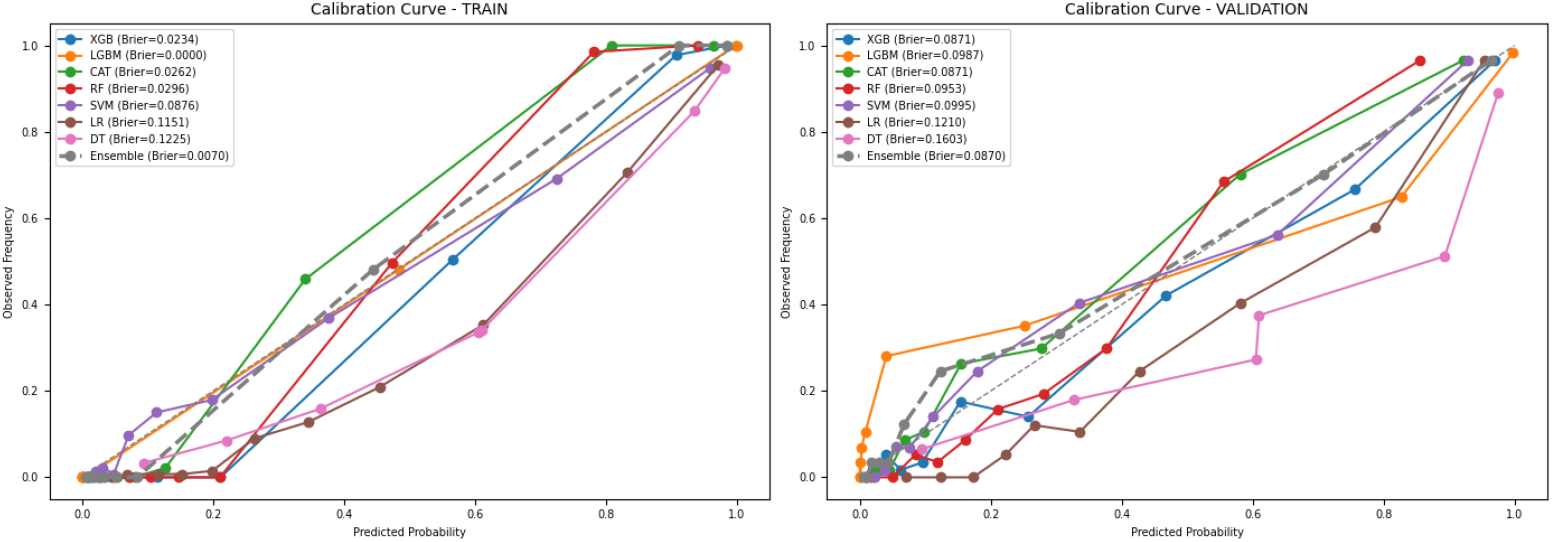
Calibration curves and Brier scores on the training and validation sets.

**Fig. 7.**
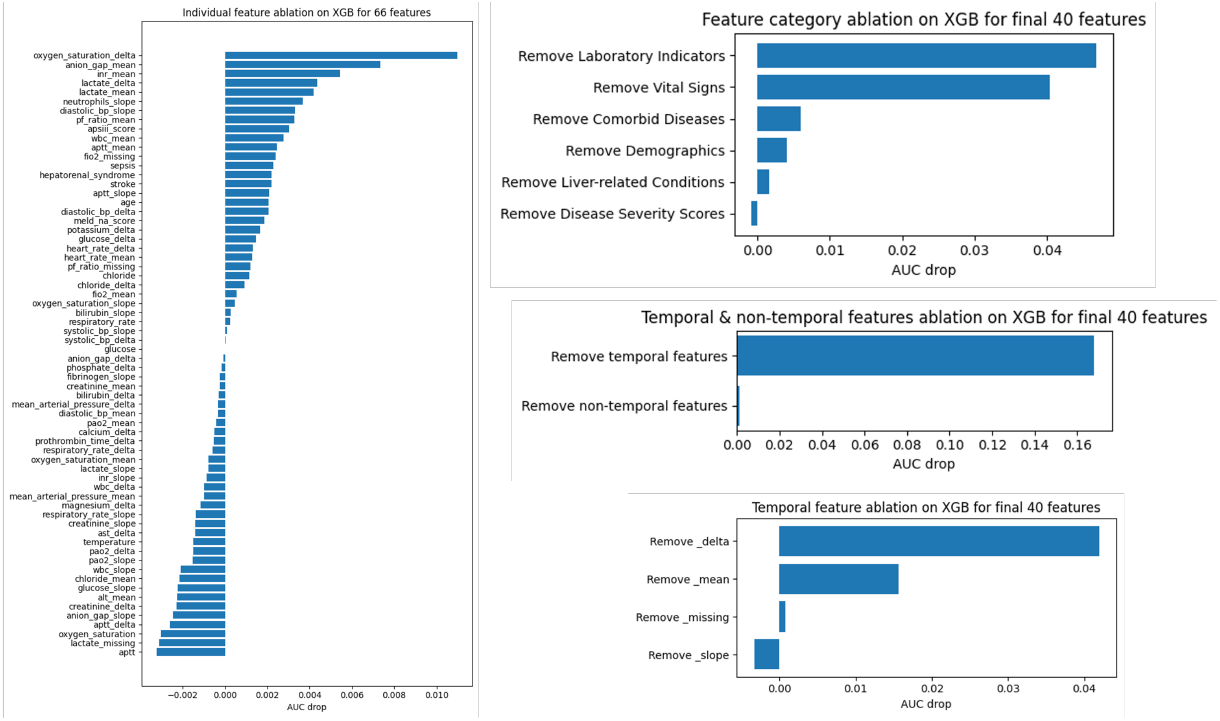
Ablation studies on the tuned XGBoost model.

### 2.10 External validation design

The frozen bundle was applied without refitting to eICU-CRD (version 2.0) [25, 26] as an independent external-validation cohort and to MIMIC-III (version 1.4) [27, 28] as a related evaluation cohort. Because MIMIC-III and MIMIC-IV originate from the same institution and cover overlapping admission periods (2008–2012), they are not fully independent. MIMIC-III was included to assess performance in a temporally related cohort, whereas eICU-CRD served as the independent external-validation dataset.

The frozen bundle comprised the 40-feature list, training-fit median imputer, refitted tree-based estimators (XGBoost, CatBoost, and LightGBM), ensemble weights, and threshold *τ* = 0.1013. External cohorts were harmonized and clinically cleaned with the same development rules, imputed with frozen training medians, and scored with the refitted ensemble as a weighted average of member probabilities; preprocessors, models, weights, and *τ* were not refit on external data.

## 3 Results

### 3.1 Baseline characteristics of the study cohort

A total of 1,907 ICU patients with alcoholic cirrhosis were included and allocated to training (*n* = 1,334, 70%) and internal validation (*n* = 573, 30%) groups with preserved 28-day mortality prevalence (24.8% in both splits). Baseline (non-temporal) characteristics are summarized in Table 4. Most variables did not differ significantly between splits. Diastolic blood pressure, P/F (PaO_2_ /FiO_2_ ) ratio, and mechanical ventilation differed at *p <* 0.05, but magnitudes were modest and not mirrored across related variables; the partitions were considered sufficiently comparable for model evaluation.

### 3.2 Final predictor set after feature selection

After filter, embedded, wrapper, and post-tuning LOO ablation, 40 predictors were retained for all final models (Table 5); 26 variables from the 66-feature RFECV set were excluded (Table 3). The final set comprised 6 static variables and 34 temporal derivatives (10 mean, 14 delta, 6 slope, 2 baseline values, and 2 missingness indicators).

### 3.3 Internal model performance

All results below used the final 40-feature predictor set, the frozen ensemble weights, and the validation-selected threshold *τ* = 0.1013 (Section 2.8; Equations 9–10). Discrimination (AUC with 95% bootstrap confidence intervals) and threshold-based classification metrics are summarized in Table 6 and Figure 3.

**Table 6.** Performance of machine learning models on the training and validation sets.

| Model | Training | Validation |  |  |  |
| --- | --- | --- | --- | --- | --- |
|  | AUC [95% CI] | AUC [95% CI] | Precision | Recall | F1-score |
| Ensemble | 1.0000 [1.0000–1.0000] | 0.9276 [0.9011–0.9507] | 0.5787 | 0.8803 | 0.6983 |
| XGBoost | 0.9983 [0.9966–0.9996] | 0.9274 [0.9004–0.9510] | 0.4481 | 0.9718 | 0.6133 |
| CatBoost | 0.9981 [0.9966–0.9992] | 0.9263 [0.8978–0.9489] | 0.5299 | 0.9366 | 0.6768 |
| LightGBM | 1.0000 [1.0000–1.0000] | 0.9258 [0.8998–0.9488] | 0.6829 | 0.7887 | 0.7320 |
| Logistic Regression | 0.9267 [0.9102–0.9409] | 0.9149 [0.8893–0.9388] | 0.2784 | 1.0000 | 0.4356 |
| Random Forest | 0.9992 [0.9982–0.9999] | 0.9146 [0.8850–0.9401] | 0.3466 | 0.9789 | 0.5120 |
| Support Vector Machine | 0.9224 [0.9051–0.9370] | 0.9095 [0.8812–0.9339] | 0.4907 | 0.9296 | 0.6423 |
| Decision Tree | 0.8892 [0.8655–0.9089] | 0.8216 [0.7758–0.8628] | 0.4006 | 0.8803 | 0.5507 |

On the training set, tree-based models showed near-perfect discrimination (ensemble and LightGBM AUC 1.0000; XGBoost 0.9983 [95% CI: 0.9966–0.9996]; CatBoost 0.9981 [95% CI: 0.9966–0.9992]), consistent with strong fit and possible overfitting. Linear models were lower but still high (logistic regression 0.9267; SVM 0.9224).

On the validation set (*n* = 573; 142 deaths), seven of eight models exceeded AUC 0.90; only the decision tree underperformed (0.8216 [95% CI: 0.7758–0.8628]). The weighted ensemble achieved the highest validation AUC (0.9276 [95% CI: 0.9011–0.9507]), followed by XGBoost (0.9274 [95% CI: 0.9004–0.9510]) and CatBoost (0.9263 [95% CI: 0.8978–0.9489]). Bootstrap confidence intervals overlapped among top gradient-boosting models, indicating modest differences in discrimination.

At *τ* = 0.1013, models differed more in precision–recall trade-offs than in AUC (Figure 4). On validation (*n* = 573; 142 deaths, 431 survivors), logistic regression and random forest achieved the highest recall (1.0000 and 0.9789) but the lowest precision (0.2784 and 0.3466), with 368 and 262 false positives, respectively— missing few deaths but flagging many survivors as high risk.

Among gradient-boosting models, LightGBM had the highest F1 (0.7320; 112 TP, 30 FN, 52 FP). CatBoost detected more deaths (133 TP, 9 FN) but with more false positives (118). The ensemble achieved the highest AUC while meeting the recall target (125 TP, 17 FN, 91 FP; precision 0.5787, recall 0.8803, F1 0.6983), with fewer false positives than logistic regression or random forest. It was therefore retained as the primary model.

### 3.4 Model interpretability (SHAP)

Model interpretability was assessed with Shapley Additive Explanations (SHAP) for the weighted tree-based ensemble (XGBoost, CatBoost, and LightGBM) on the internal validation set (Figure 5). The bar plot ranks predictors by mean absolute SHAP value; the beeswarm plot shows how each feature shifts predicted mortality risk (red = higher feature values, blue = lower).

APS III score was the strongest contributor, with higher scores increasing predicted mortality. Anion gap (mean and delta) ranked next among laboratory summaries; higher values increased risk. Among temporal vital-sign features, oxygen saturation (delta) was highly ranked, with lower delta values associated with higher risk, consistent with worsening oxygenation during the ICU stay. International normalized ratio (mean), lactate (mean), and activated partial thromboplastin time (mean) also increased risk at higher values. Lower diastolic blood pressure (delta) and lower P/F ratio (mean) were associated with higher mortality. White blood cell count (mean and delta), creatinine (mean), and heart rate (delta and mean) captured dynamic physiology in directions consistent with clinical deterioration. Sepsis, stroke, and older age also increased predicted risk.

Overall, the ensemble relied on clinically interpretable severity scores (APS III, MELD-Na), laboratory trajectories and summaries, vital-sign derivatives, and comorbidities. Temporal features featured prominently among the top contributors—often alongside corresponding mean or baseline measurements— supporting the value of trajectory-based predictors beyond static admission values.

### 3.5 Calibration

Calibration was evaluated with reliability curves and the Brier score (lower = better probabilistic accuracy), as shown in Figure 6. On the training set, gradient-boosting models and the ensemble had very low Brier scores (e.g., ensemble 0.0070; LightGBM 0.0000), with curves near the diagonal, consistent with strong fit and possible overfitting.

On the validation set, the ensemble had the lowest Brier score (0.0870), followed by XGBoost and CatBoost (both 0.0871). LightGBM (0.0987), random forest (0.0953), and SVM (0.0995) were modestly higher; logistic regression (0.1210) and the decision tree (0.1603) showed the poorest calibration. Ensemble and gradient-boosting curves tracked the diagonal more closely than logistic regression and the decision tree, indicating more reliable probability estimates for the primary model.

### 3.6 Ablation analysis

Ablation results for the tuned XGBoost model are shown in Figure 7. LOO ablation used the 66-feature set (baseline validation AUC 0.9094); group and temporal ablations used the final 40-feature set (baseline validation AUC 0.9274). All outcomes are reported as validation AUC changes and are independent of the classification threshold.

The largest individual-feature decreases were after removing oxygen saturation (delta; drop 0.0110), anion gap (mean; 0.0073), international normalized ratio (mean; 0.0054), lactate (delta; 0.0044), lactate (mean; 0.0042), neutrophils (slope; 0.0037), and APS III score (0.0030). Twenty-six features showed **Δ**AUC ≤ 0 and were excluded from the final set.

At the group level, removing laboratory indicators caused the largest loss (drop 0.0468; AUC 0.8806), followed by vital signs (drop 0.0404; AUC 0.8870). Smaller decreases were observed for comorbid diseases (0.0059), demographics (0.0041), and liver-related conditions (0.0016); removing severity scores had minimal impact (drop −0.0008).

Removing all temporal features reduced validation AUC to 0.7595 (drop 0.1680), whereas removing non-temporal features reduced AUC to 0.9264 (drop 0.0011). Among temporal representations, delta features had the greatest influence (drop 0.0419), followed by mean features (0.0157) and missingness indicators (0.0008); removing slope features slightly increased AUC (drop −0.0033).

### 3.7 External validation

The frozen development bundle was applied without refitting to eICU-CRD (version 2.0; *n* = 481, 19.1% mortality) and MIMIC-III (version 1.4; *n* = 791, 28.1% mortality). Performance at threshold 0.1013 is summarized in Table 7.

**Table 7.** External validation performance across datasets (threshold = 0.1013).

| Model | eICU-CRD |  |  |  | MIMIC-III |  |  |  |
| --- | --- | --- | --- | --- | --- | --- | --- | --- |
|  | AUC [95% CI] | Prec. | Rec. | F1 | AUC [95% CI] | Prec. | Rec. | F1 |
| Ensemble | 0.9347 [0.8963–0.9650] | 0.7865 | 0.7609 | 0.7735 | 0.9071 [0.8784–0.9304] | 0.7425 | 0.7793 | 0.7604 |
| XGBoost | 0.9346 [0.8956–0.9653] | 0.6500 | 0.8478 | 0.7358 | 0.9056 [0.8773–0.9291] | 0.5710 | 0.8874 | 0.6949 |
| CatBoost | 0.9338 [0.8958–0.9630] | 0.6786 | 0.8261 | 0.7451 | 0.9038 [0.8740–0.9283] | 0.6619 | 0.8378 | 0.7396 |
| LightGBM | 0.9245 [0.8825–0.9585] | 0.9167 | 0.7174 | 0.8049 | 0.9008 [0.8724–0.9258] | 0.8201 | 0.6982 | 0.7543 |
| Random Forest | 0.9207 [0.8792–0.9563] | 0.3836 | 0.9130 | 0.5402 | 0.8988 [0.8676–0.9250] | 0.4764 | 0.9099 | 0.6254 |
| Support Vector Machine | 0.9177 [0.8747–0.9543] | 0.8000 | 0.6522 | 0.7186 | 0.8802 [0.8502–0.9076] | 0.3182 | 0.9775 | 0.4801 |
| Logistic Regression | 0.9171 [0.8749–0.9527] | 0.3816 | 0.9457 | 0.5438 | 0.8778 [0.8487–0.9052] | 0.5974 | 0.8153 | 0.6895 |
| Decision Tree | 0.8126 [0.7572–0.8596] | 0.5328 | 0.7065 | 0.6075 | 0.8147 [0.7761–0.8509] | 0.4844 | 0.8378 | 0.6139 |

On eICU-CRD, the ensemble achieved AUC 0.9347 (95% CI: 0.8963–0.9650), similar to XGBoost (0.9346 [95% CI: 0.8956–0.9653]), with precision 0.7865, recall 0.7609, and F1 0.7735. On MIMIC-III, the ensemble achieved AUC 0.9071 (95% CI: 0.8784–0.9304), comparable to XGBoost (0.9056 [95% CI: 0.8773–0.9291]), with precision 0.7425, recall 0.7793, and F1 0.7604. Gradient-boosting models outperformed the decision tree on both cohorts, and bootstrap confidence intervals overlapped among top models. Discrimination remained strong despite differences in cohort size, mortality prevalence, and data source, supporting transportability of the frozen pipeline; eICU-CRD provides the more rigorous independent external test, whereas MIMIC-III reflects performance in a related institutional cohort.

## 4 Discussion

### 4.1 Summary of findings

This study developed a trajectory-informed machine learning framework for 28-day all-cause mortality in ICU patients with alcoholic cirrhosis using MIMIC-IV. A multi-stage pipeline reduced 208 engineered predictors to a compact 40-feature set, and seven learners were benchmarked alongside a weighted gradient-boosting ensemble. On internal validation, the ensemble achieved the strongest discrimination (AUC 0.9276 [95% CI: 0.9011–0.9507]) and the lowest Brier score (0.0870), while meeting the prespecified recall-oriented classification target at *τ* = 0.1013. At that threshold, it also offered a more balanced precision–recall trade-off than logistic regression or random forest, which achieved very high recall only by generating large numbers of false-positive alerts among survivors. Performance remained strong on eICU-CRD (AUC 0.9347 [95% CI: 0.8963–0.9650]) and on the related MIMIC-III cohort (AUC 0.9071 [95% CI: 0.8784–0.9304]) when the development bundle was applied without refitting; eICU-CRD provides the more rigorous independent external test, whereas MIMIC-III reflects performance in an overlapping institutional cohort.

Three findings are clinically and methodologically notable. First, temporal ablation showed that dynamic representations dominated model contribution: removing all temporal features reduced validation AUC by 0.1680, whereas removing non-temporal features reduced AUC by only 0.0011. This supports encoding physiological change, not only static admission values, in ICU mortality modeling. Second, group ablation indicated that laboratory and vital-sign domains drove most of the signal, with smaller decreases after removing comorbidity, demographic, or liver-specific groups, consistent with multisystem deterioration in decompensated cirrhosis. Third, SHAP analysis showed clinically plausible drivers: APS III score, anion gap (mean and delta), oxygen saturation (delta), lactate (mean), international normalized ratio (mean), sepsis, and P/F ratio (mean), with temporal derivatives prominent among top contributors. Additional signal came from coagulation markers, white blood cell trajectories, blood pressure and heart rate derivatives, and age; many top predictors appeared alongside corresponding mean or baseline measurements, suggesting integration of evolving physiology with static severity rather than reliance on admission values alone.

Tree-based gradient boosting was well suited to this tabular feature space because it can model non-linear effects and interactions among laboratories, vitals, and severity measures; the weighted ensemble further combined complementary boosting implementations and improved stability relative to any single model. Together, these results suggest that the ensemble captures severity, organ dysfunction, and evolving physiology in directions aligned with bedside intuition, while remaining discriminative, reasonably well calibrated, and transportable under frozen external evaluation.

### 4.2 Comparison with prior studies

Prior work in this population has relied heavily on conventional scores such as MELD, APACHE II, and SOFA, which use limited static inputs and modest discrimination. A prior MIMIC-IV study [13] showed that machine learning could outperform traditional scores for alcoholic cirrhosis mortality, but relied primarily on static features. The present study extends that line of work by systematically engineering mean, delta, slope, baseline, and missingness representations from longitudinal vitals and laboratories, then selecting predictors through filter, embedded, wrapper, and post-tuning ablation stages. Removing all temporal features reduced validation AUC by 0.1680, a larger effect than incremental differences among top gradient-boosting algorithms and a meaningful advance over static feature approaches in this setting.

Methodologically, the study contributes a reproducible development workflow with explicit leakage control, multi-model benchmarking, calibration assessment, SHAP interpretability, and frozen external evaluation. Benchmarking showed that tree-based gradient-boosting models outperformed linear baselines, while the weighted ensemble provided the best overall combination of discrimination and calibration on internal validation. eICU-CRD provides the more rigorous independent external test; MIMIC-III should be interpreted as a related institutional cohort because of patient overlap with MIMIC-IV. Together, these elements suggest that feature representation and evaluation design may be as important as model class selection for ICU mortality prediction in alcoholic cirrhosis.

Relative to conventional severity scores, the framework uses a broader nonlinear feature space while retaining clinically recognizable inputs such as APS III, lactate, coagulation tests, oxygenation measures, and sepsis. Because these predictors are routinely available in ICU electronic health records, the model may support early risk stratification after local threshold and calibration review; strong retrospective performance alone does not establish clinical utility.

### 4.3 Limitations and future work

This study has several limitations. The analysis used retrospective electronic health record data, which are subject to missingness, documentation bias, coding variation, and measurement heterogeneity across time and units; although missing values were handled with training-fit median imputation and consistently applied frozen imputation externally, informative missingness could not be fully modeled. Temporal predictors were aggregated summaries over the ICU observation window rather than continuous irregular time series, so brief events such as acute hemodynamic decline, transient hypoxemia, or rapid metabolic shifts may be attenuated. External evaluation was restricted to U.S. tertiary-care databases: MIMIC-III overlaps with MIMIC-IV in institution and period and shares patients with the development cohort, whereas eICU-CRD is the stronger independent test; outcome prevalence also differed across cohorts (MIMIC-IV 24.8%, eICU-CRD 19.1%, MIMIC-III 28.1%), underscoring the need for site-specific validation, recalibration, and threshold review. Post-tuning leave-one-out feature exclusion used internal validation AUC and may introduce modest optimism relative to a fully nested selection procedure. Finally, all metrics reflect retrospective discrimination and calibration and do not demonstrate that the model improves triage, monitoring, resource allocation, or patient outcomes in real clinical use.

Future work should evaluate irregular time-series methods better suited to asynchronous clinical measurements, multi-center training approaches such as federated learning to improve geographic and demographic coverage without sharing raw records, and prospective implementation studies with embedded decision-support workflows. Clinical value should be assessed not only by AUC, but by whether predicted risk changes clinician behavior and improves care processes and outcomes in practice.

## 5 Conclusions

This study demonstrates that a machine learning framework combining temporal feature engineering with weighted gradient-boosting ensemble learning can accurately predict 28-day all-cause mortality in ICU patients with alcoholic cirrhosis. By encoding longitudinal vitals and laboratories as trajectory-based summaries rather than static admission values alone, the approach captured dynamic physiological deterioration poorly represented by conventional risk tools. A multi-stage selection pipeline yielded a compact 40-feature predictor set, and the final ensemble achieved strong internal discrimination and calibration (validation AUC 0.9276 [95% CI: 0.9011–0.9507]; Brier score 0.0870), with preserved performance on eICU-CRD (AUC 0.9347 [95% CI: 0.8963–0.9650]) and related MIMIC-III evaluation (AUC 0.9071 [95% CI: 0.8784–0.9304]) under a frozen development bundle. Ablation and SHAP analyses indicated that temporal change features and laboratory–vital domains drove most of the signal, with APS III, anion gap, oxygen saturation trajectories, lactate, coagulation markers, sepsis, and P/F ratio among the leading contributors—findings that are clinically plausible and consistent with multisystem deterioration in decompensated cirrhosis. These results support trajectory-informed risk stratification in this high-risk population; nonetheless, the framework remains a retrospective demonstration, and prospective, multi-center validation with site-specific threshold and calibration adjustment is required before clinical deployment.

## Data Availability

The datasets analyzed during the current study are publicly available through PhysioNet under data use agreements. MIMIC-IV (version 3.1) is available at https://doi.org/10.13026/kpb9-mt58, MIMIC-III (version 1.4) is available at https://doi.org/10.13026/C2XW26, and eICU-CRD (version 2.0) is available at https://doi.org/10.13026/C2WM1R. Access to these databases requires credentialed registration and completion of the required training. The code supporting the findings of this study is available from the corresponding author upon reasonable request.

https://doi.org/10.13026/C2XW26

https://doi.org/10.13026/C2WM1R

## Declarations

### Ethics approval and consent to participate

This study is a secondary analysis of de-identified, publicly available data and didn not require additional institutional review board approval. Access to MIMIC-IV, MIMIC-III, and eICU-CRD was granted after completion of the required human-subjects training and execution of the PhysioNet credentialed-access data use agreements. The original creation of these databases was approved by the relevant institutional review boards (for MIMIC, the Beth Israel Deaconess Medical Center and the Massachusetts Institute of Technology), with the requirement for individual patient consent waived because all records are de-identified. The study followed the principles of the Declaration of Helsinki.

### Consent for publication

Not applicable.

### Availability of data and materials

The datasets analyzed during the current study are publicly available through PhysioNet under data use agreements. MIMIC-IV (version 3.1) is available at https://doi.org/10.13026/kpb9-mt58, MIMIC-III (version 1.4) is available at https://doi.org/10.13026/C2XW26, and eICU-CRD (version 2.0) is available at https://doi.org/10.13026/C2WM1R. Access to these databases requires credentialed registration and completion of the required training. The code used for data preprocessing, feature engineering, model development, and analysis is available from the corresponding author upon reasonable request.

### Authors’ contributions

JS conceived the study, developed the research idea, curated the data, designed the methodology, performed the validation, prepared all figures and tables, wrote the original manuscript draft, and revised the manuscript. MH contributed to methodology and manuscript review and editing. NK contributed to methodology and manuscript review and editing. SP contributed to methodology and manuscript review and editing. SVC contributed to methodology and manuscript review and editing. MP supervised the study and contributed to manuscript review and editing. KA offered expert clinical advice. All authors reviewed and approved the final manuscript.

## Acknowledgements

Not applicable.

